# Genetic analysis of non-familial early-onset Alzheimer’s disease patients

**DOI:** 10.64898/2026.11.20.26367760

**Authors:** F Ecca, L Nicholson, M Naymik, A Bonfitto, IS Piras, RJ Caselli, BK Woodruff, TG Beach, GE Serrano, MJ Huentelman, CR Padilla

**Affiliations:** Division of Early Detection and Prevention, Translational Genomics Research Institute, Phoenix, AZ, USA; Department of Neurology, Mayo Clinic, Scottsdale, AZ, USA; Banner Sun Health Research Institute, Sun City, AZ, USA; Baylor University Medical Center, Dallas, TX, USA

**Keywords:** early-onset Alzheimer’s Disease (EOAD), late-onset AD (LOAD), polygenic risk score (PRS), APOE4, rare variants

## Abstract

Early-onset Alzheimer’s disease (EOAD), defined by symptom onset before age 65, can be either familial or sporadic and account for ∼5% of all Alzheimer’s disease (AD) cases, with only ∼10% showing Mendelian inheritance. In this study, we compare the genetic architecture of four cohorts comprising 87 sporadic EOAD cases, 1,018 late-onset AD (LOAD) cases, 585 non-demented control (TGEN2-AD and TGEN2-ND cohorts), and 3,487 non-demented controls from the AD Sequencing Project (ADSP-CTL).

We observed a significant enrichment of the APOE ε4 allele in EOAD cohort compared to controls (52.2% vs 20.1%; *p* = 1.54e-10), although its frequency was lower than in the LOAD cohort (64.5%; *p* = 0.0298). As expected, APOE ε4 was strongly enriched in the LOAD group relative to controls (*p* = 1.23e-63). Sporadic EOAD genome-wide association (GWAS) analysis and sequence kernel association tests (SKAT) confirmed strong association signals in the *APOE* locus, including the genes *APOE, APOC1, NECTIN2*, and *TOMM40*. AD-related Polygenic Risk Score (PRS) showed that EOAD affected patients carried an intermediate genetic risk (mean PRS = 40.1), significantly lower than LOAD (mean PRS = 57.5; *p* = 6.7e-05) but higher than controls (mean PRS = 25.1; *p* = 6.1e-04). Notably, rare variants in autosomal dominant AD genes (*APP, PSEN1, PSEN2*) and in previously reported EOAD/LOAD modifier genes where not enriched in sporadic EOAD cohort.

These results suggest, despite earlier onset, sporadic EOAD is not driven by a higher polygenic burden or rare high-impact variants. Instead, its genetic architecture appears to overlap more closely with LOAD, supporting the hypothesis that EOAD may represent an early and more severe manifestation of LOAD, or a genetically distinct subtype. Unidentified risk factors, such as epigenetic modifications or complex structural variants, may contribute to EOAD genetic risk and warrant further investigation to better elucidate the complex genetic architecture of Alzheimer’s disease.

## INTRODUCTION

Alzheimer’s disease (AD) is the leading cause of dementia, presenting a growing healthcare challenge with approximately 6.7 million diagnosed cases in the United States^1^. Pathological hallmarks include the accumulation of abnormal protein aggregates, including beta-amyloid (Aβ) in senile plaques and hyperphosphorylated tau in neurofibrillary tangles (NFTs) driven by a complex interplay between genetic, environmental, and other factors. AD can be classified into two primary types: early-onset and late-onset AD. Early-onset AD accounts for 5-10% of all cases and is marked by symptom onset before the age of 65^2,3^. Individuals with a familial history of early-onset AD typically exhibit causal genetic mutations in one of three key genes, *Amyloid Precursor Protein* (*APP*) or the *Presenilins* (*PSEN1* and *PSEN2*)^3–5^. However, about 90% of early-onset cases arise sporadically where patients often lack common heritable high-risk genetic variants^3,4^.

To better understand the contribution of common genetic variation to AD susceptibility, polygenic risk scores (PRS) have been developed. These scores aggregate the effects of multiple common variants identified through genome-wide association studies (GWAS). Nevertheless, PRS largely overlook the potential role of rare and private variants, which have already been implicated in AD risk and pathogenesis. In particular, private variants in genes with common variant association signals such as *TREM2, SORL1*, and *ABCA7*, among others, are increasingly recognized as potential contributors to disease susceptibility, even in the absence of high-penetrance mutations^2,6^.

In the era of precision medicine, the significant impact of rare genomic variants (minor allele frequency <1%), including single nucleotide polymorphisms (SNPs), copy number variations, variable number tandem repeats, and structural variations, whether *de novo* or inherited, germline or somatic, has been emphasized through extensive studies utilizing large genomic datasets^7^. These rare variants are implicated in a variety of human diseases, spanning from monogenic to multifactorial disorders, including neurodegenerative diseases, cancer, and various physiological conditions^3,7–14^. While early investigations focused on the effects of rare variants in coding regions, research has progressed to employing whole exome sequencing (WES) and ultimately whole genome sequencing (WGS) due to decreasing costs. Despite these advancements and increased cohort sizes, analyses have predominantly concentrated on protein-coding effects, limiting our ability to decipher the intricate regulation of human genes and their contributions to pathological phenotypes and risk. Additionally, many small case-control studies have focused on a limited number of genes or putatively high-impact variants, making it difficult to replicate the identification of pathogenic variants in early-onset AD cases without a familial history, as well as in other forms of the disease.

To further explore the presence or absence of heritable common traits in patients affected by sporadic EOAD and the significant influence of rare variants, we conducted a GWAS and Sequence Kernel Association Test (SKAT) on 87 sporadic EOAD patients, 1,018 LOAD and 4,072 not demented control samples. We additionally calculated an AD-related PRS using genes associated with familial and sporadic AD. Our findings revealed multiple significant associations with the *APOE* locus (p < 5.0e-08) for both common and rare variants, and confirm a similar genomic risk between sporadic EOAD and LOAD compared with control population where LOAD show a significant higher risk^15^. A similar trend was observed in the distribution of the APOE ε4 allele, where both EOAD and LOAD cohorts were enriched for the risk allele when compared with control group (p = 1.54e-10, and p = 1.23e-63 respectively), with LOAD showing the strongest enrichment. However, in contrast to some existing literature highlighting an enrichment of rare variants in the dominant autosomal genes, as well in modifier genes for EOAD, we were unable to identify significant differences from the control cohort or in other genes previously associated with LOAD^16^.

## METHODS

### Inclusion criteria and data analysis

For the remainder of this manuscript, the simplified acronym EOAD will be used to refer to sporadic (e.g., non-familial) early-onset AD, while LOAD will be used to refer to late-onset AD. Detail regarding demographic and clinical characteristics of the cohorts are summarized in ***Table 1***. The EOAD group included 88 patients (females: n = 51; males: n = 37), from which one sample was discarded during quality control (see results for details). Patients were classified as sporadic based on clinical history and hospital records, this was subsequently supported by WES data confirming the absence of pathogenic variants in *APP, PSEN1*, and *PSEN2*. The TGEN2-ND cohort includes 585 non-demented controls individuals, whereas the TGEN2-AD cohort includes 1,014 neuropathologically confirmed LOAD patients. DNA samples were extracted from whole blood using the QIAamp DNA Blood Midi Kits (Qiagen, USA) and analyzed with WES, using the Illumina technology. Libraries were prepared with either the TruSeq 62Mb Exome Enrichment Kit or the Nextera 62Mb Rapid Capture Expanded Exome Kit (Illumina, Inc., San Diego, CA, USA), following manufacturers’ protocols. Exome libraries were sequenced by 100-bp paired-end sequencing on a HiSeq 2500 System (Illumina, Inc., San Diego, CA, USA). Control data were obtained from the Alzheimer’s disease sequencing project (ADSP) and downloaded from dbGAP (accession number: phs000572.v.p4) following an approved request. In the remainder of the manuscript, we refer to this cohort as ADSP-CTL. We selected only neurological controls who were 80 or older (n = 3,501; females: n = 2,046; males: n = 1,455). After sex check analysis, we removed 14 samples from the ADSP-CTL group (see results), obtaining a final sample size of 3,487. FASTQ files for the EOAD group and ADSP-CTL were aligned to the human reference genome *GRCh38* using the *parabricks* pipeline *v4*.*0*.*1*. Joint genotyping was then conducted using *GLnexus v1*.*4*.*1*. The resulting VCF file was filtered, excluding variants with quality score (QUAL) < 300, depth (DP) < 10, genotype quality (GQ) < 20, SNPs located within 10 bp of insertion/deletions (INDELS), and allelic balance for heterozygous genotypes outside the range of 0.2 to 0.8. Additionally, we retained only biallelic SNPs, excluding INDELs. Subsequently, the file was converted into PLINK format to conduct sex-mismatch analysis and further SNPs and sample filtering^17^. The missing sex for four EOAD samples was imputed. We removed SNPs with a genotyping rate < 95%, minimum allele frequency (MAF) < 0.1%, and those not meeting Hardy-Weinberg Equilibrium with p < 1.0e-06. Samples with a genotyping rate lower than 20% were also excluded. The dataset underwent imputation using the Michigan Imputation Server 2 (MIS2) with the 1000 Genomes v3 30X reference panel, and only including imputed variants with r^2^ ≥ 0.3. The dataset was then re-filtered for MAF, HWE, SNPs and sample genotyping rate. Duplicate samples were assessed using the pi-hat metric in PLINK, with duplicates defined as those with a pihat ≥ 0.7. Principal component analysis was used to identify outlier-based samples above the cutoff of ± 6 Standard Deviation across the top 7 principal components, representing more than 50% of the total variance.

**Table 1.**
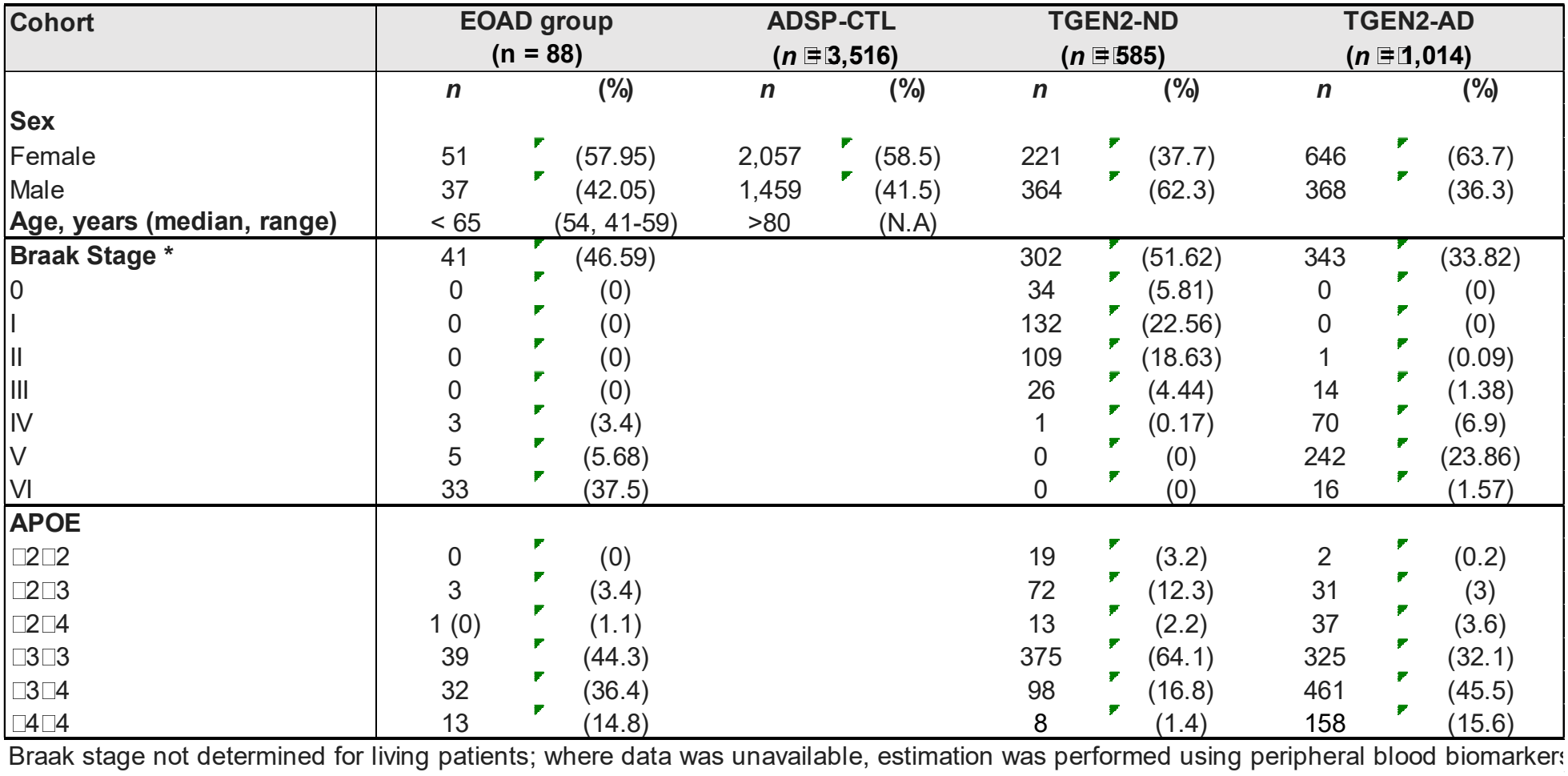
Demographic and clinical characteristic of AD patients and control samples enrolled in the study.

| Cohort | EOAD group<br>(n = 88) |  | ADSP-CTL<br>(*n* =□3,516) |  | TGEN2-ND<br>(*n* =□585) |  | TGEN2-AD<br>(*n* =□1,014) |  |
|---|---|---|---|---|---|---|---|---|
|  | ***n*** | **(%)** | ***n*** | **(%)** | ***n*** | **(%)** | ***n*** | **(%)** |
| **Sex** |  |  |  |  |  |  |  |  |
| Female | 51 | (57.95) | 2,057 | (58.5) | 221 | (37.7) | 646 | (63.7) |
| Male | 37 | (42.05) | 1,459 | (41.5) | 364 | (62.3) | 368 | (36.3) |
| **Age, years (median, range)** | < 65 | (54, 41-59) | >80 | (N.A) |  |  |  |  |
| **Braak Stage \*** | 41 | (46.59) |  |  | 302 | (51.62) | 343 | (33.82) |
| 0 | 0 | (0) |  |  | 34 | (5.81) | 0 | (0) |
| I | 0 | (0) |  |  | 132 | (22.56) | 0 | (0) |
| II | 0 | (0) |  |  | 109 | (18.63) | 1 | (0.09) |
| III | 0 | (0) |  |  | 26 | (4.44) | 14 | (1.38) |
| IV | 3 | (3.4) |  |  | 1 | (0.17) | 70 | (6.9) |
| V | 5 | (5.68) |  |  | 0 | (0) | 242 | (23.86) |
| VI | 33 | (37.5) |  |  | 0 | (0) | 16 | (1.57) |
| **APOE** |  |  |  |  |  |  |  |  |
| □2□2 | 0 | (0) |  |  | 19 | (3.2) | 2 | (0.2) |
| □2□3 | 3 | (3.4) |  |  | 72 | (12.3) | 31 | (3) |
| □2□4 | 1 (0) | (1.1) |  |  | 13 | (2.2) | 37 | (3.6) |
| □3□3 | 39 | (44.3) |  |  | 375 | (64.1) | 325 | (32.1) |
| □3□4 | 32 | (36.4) |  |  | 98 | (16.8) | 461 | (45.5) |
| □4□4 | 13 | (14.8) |  |  | 8 | (1.4) | 158 | (15.6) |

Braak stage not determined for living patients; where data was unavailable, estimation was performed using peripheral blood biomarkers

### Rare variant analysis

Variant annotation was performed using the Ensembl Variant Effect Predictor (VEP), based on GENCODE v34 gene models. Variants were filtered to retain only those with potential functional impact, including exonic, untranslated regions (UTRs), and splice donor and acceptor sites. To prioritize putatively deleterious rare variants, we selected those with a Combined Annotation Dependent Depletion (CADD) score > 23 and a minor allele frequency (MAF) < 1%. A curated list of 83 genes previously associated with EOAD or LOAD, based on prior sequencing and GWAS studies, was used for targeted analysis (see Supplementary Table 1).

### GWAS and SKAT analysis

After these filtering steps, we generated a dataset for the SKAT analysis with MAF ≥ 0.1% and another for the genome-wide association study (GWAS) with MAF ≥ 5% GWAS was conducted using the *BOLT-LMM* algorithm, adjusting for sex and the top 10 principal components^18^. *BOLT-LMM* algorithm employs a linear mixed model assuming a Bayesian mixture-of-normals prior for the random effect attributed to SNPs other than the one being tested. This increases power compared to previous mixed-model association methods such as *GEMMA, FastLMM* and *EMMAX*, while efficiently adjusting for population stratification and relatedness. SNPs with p < 5.0e-08 were considered significant. The p-value distribution was assessed with BOLT-LMM software using the lambda genomic control (λ-GC) inflation factor, with λ-GC < 1.1 indicating the absence of significant inflation. The SKAT test at the gene level was conducted using GENESIS R-package^19^. SNPs were assigned to genes defined by regions with overlapping transcripts from the same gene, considering only regions with 5 or more SNPs. The results were adjusted with the Bonferroni method, accounting for the number of regions tested. P-value distribution was assessed using λ, with λ < 1.1 indicating the absence of significant inflation.

### Statistical analysis and AD-Polygenic risk score

To compare the distribution of the APOE ε4 allele between groups, a chi-squared test for equality of proportions was performed using the prop.test function in R. The AD Polygenic Risk Score (AD-PRS) was calculated as described by Marees et al.^20^ using the SNPs from Lambert et al.^21^ across three cohorts: EOAD, neuropathologically confirmed sporadic AD (TGEN2-AD) and non-dement controls (TGEN2-ND), which was obtained and previously described by Corneveaux et al.^22^. We selected SNPs with False Discovery Rate (FDR) less than 0.05 (n = 3,799), and mapped to the Human Genome Reference Hg38. The number of alleles present in each individual were counted and then multiplied by their corresponding GWAS b-value and finally summed across all SNPs for each individual’s PRS. A Welch two-sample t-test was used to compare PRS scores between groups. All data processing and statistical analyses were performed in R.

### RESULTS

### Significant associations with the *APOE* locus and similar genomic risk (*APOC1, NECTIN2*, and *TOMM40)* were found from GWAS analysis between sporadic EOAD and sporadic LOAD

The clinical and demographic characteristics of the study populations are presented in ***Table 1***. After SNPs quality controls, we obtained 87 EOAD, 3,487 ADSP-CTL, 1,018 TGEN2-AD and 585 TGEN2-ND. To assess the impact of *APOE* alleles in the risk of developing different forms of AD in our cohorts, we calculated the distribution of the three main alleles: *APOE2* (ε2), *APOE3* (ε3), and *APOE4* (ε4) (***Table 1***). The APOE ε4 allele, the strongest known genetic risk factor for neurodegenerative disease, shows marked variability in frequency across ancestries and geographic ranging from ∼15%-25% in the general population, about 40% in Central Africa populations, and varying within Europe and Asia along regional gradients^23–26^. In our cohorts, ε4 frequency increased from 20.1% in the control group (TGEN2-ND) to 52.2% in EOAD affected patients and reached 64.5% in the LOAD cohort (TGEN2-AD) (***Figure 1***). The ε4 allele was significantly more frequent in the EOAD cohort compared to controls (p = 1.54e-10), but significantly less frequent than in the LOAD cohort (p = 0.0298). As expected, the LOAD group was significantly enriched for APOE ε4 (p = 1.23e-63) versus the controls. Despite this overall enrichment of the ε4 allele among AD cases, no significant difference was observed in the distribution of APOE ε4 homozygotes between EOAD and LOAD (p-value = 0.8406).

**Figure 1:**
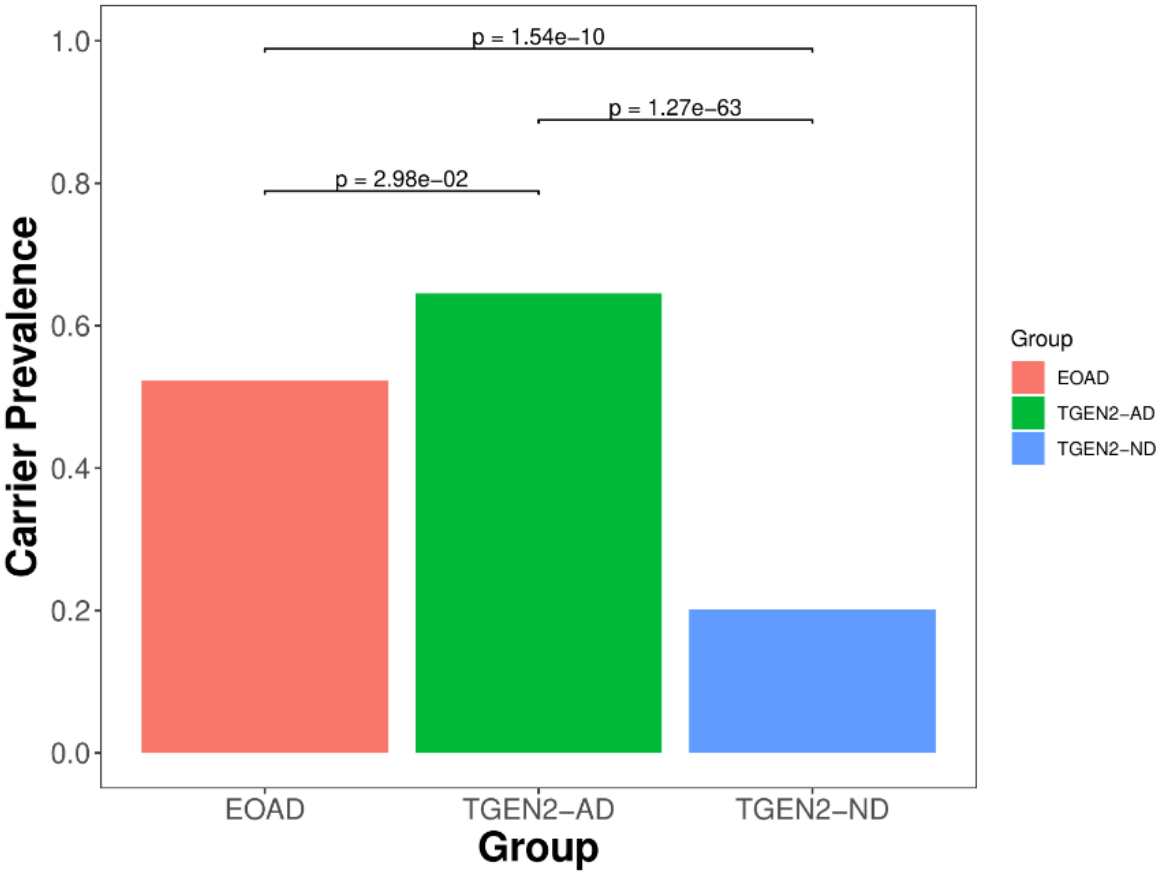
APOE4 allele distribution across the cohorts.

After joint genotype calling and preliminary VCF filtering, we identified 683,359 SNPs across 88 EOAD and 3501 ADSP-CTL samples. Following sex mismatch analysis, 14 ADSP-CTL samples were excluded. The missing sex for four EOAD samples was imputed using PLINK. We decided to not remove those 4 samples to not decrease the already limited sample size for the EOAD group. One EOAD sample was removed for low genotyping rate (< 80%), resulting in a final dataset comprising 87 EOAD and 3,487 CTL. Post-SNP filtering, we retained 94,077 variants with MAF ≥ 0.1% (total genotyping rate: 99.4%), and subsequent imputation with the MIS2 expanded the dataset to 12,887,939 variants. Further quality control steps were conducted to detect potential outliers, but none were identified. We then created two datasets: the GWAS dataset included SNPs with MAF ≥ 5% (totaling 3,263,868 SNPs), while the SKAT dataset also encompassed both rare and common SNPs with MAF ≥ 0.1% (totaling: 7,532,700 SNPs). The PCA plot of the final dataset is shown in ***Figure S2***.

The GWAS, adjusted for sex and 10 PCs, yielded 31 significant SNPs (p < 5.0e-08), all located on chromosome 19 and encompassing the genes *APOE, APOC1, NECTIN2*, and *TOMM40*. The most significant SNP was rs429358 at chr19:44908684 (p = 1.7E-33) which, together with rs7412, defines the APOE ε2/ε3/ε4 isoforms (***Table 2; Figure 2)***. SKAT analysis was conducted on 30,031 regions (including 5 SNPs or more), setting the significance level at p < 1.67e-06 and adjusting for sex and the top 10 principal components, four genes, also detected in the GWAS, achieved the gene-wide significance: *NECTIN2, APOE, APOC1* and *TOMM40* (***Figure 3; Table 3***).

**Table 2.**
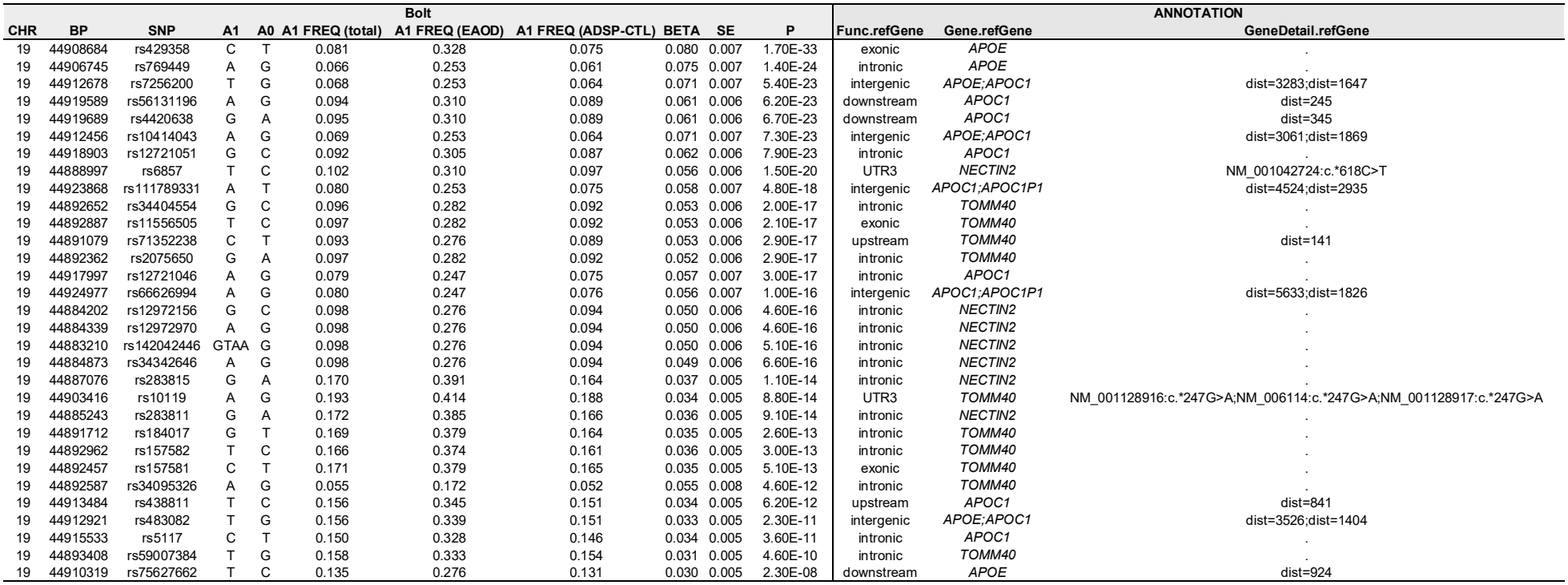
Significant SNPs (p < 5.0E-08) obtained in the GWAS comparison EAOD (n = 87) and ADSP-CTL (n = 3487). A1: effect allele; A2; other allele.

| Bolt |  |  |  |  |  |  |  |  |  |  | ANNOTATION |  |  |
|---|---|---|---|---|---|---|---|---|---|---|---|---|---|
| CHR | BP | SNP | A1 | A0 | A1 FREQ (total) | A1 FREQ (EAOD) | A1 FREQ (ADSP-CTL) | BETA | SE | P | Func.refGene | Gene.refGene | GeneDetail.refGene |
| 19 | 44908684 | rs429358 | C | T | 0.081 | 0.328 | 0.075 | 0.080 | 0.007 | 1.70E-33 | exonic | *APOE* | . |
| 19 | 44906745 | rs769449 | A | G | 0.066 | 0.253 | 0.061 | 0.075 | 0.007 | 1.40E-24 | intronic | *APOE* | . |
| 19 | 44912678 | rs7256200 | T | G | 0.068 | 0.253 | 0.064 | 0.071 | 0.007 | 5.40E-23 | intergenic | *APOE;APOC1* | dist=3283;dist=1647 |
| 19 | 44919589 | rs56131196 | A | G | 0.094 | 0.310 | 0.089 | 0.061 | 0.006 | 6.20E-23 | downstream | *APOC1* | dist=245 |
| 19 | 44919689 | rs4420638 | G | A | 0.095 | 0.310 | 0.089 | 0.061 | 0.006 | 6.70E-23 | downstream | *APOC1* | dist=345 |
| 19 | 44912456 | rs10414043 | A | G | 0.069 | 0.253 | 0.064 | 0.071 | 0.007 | 7.30E-23 | intergenic | *APOE;APOC1* | dist=3061;dist=1869 |
| 19 | 44918903 | rs12721051 | G | C | 0.092 | 0.305 | 0.087 | 0.062 | 0.006 | 7.90E-23 | intronic | *APOC1* | . |
| 19 | 44888997 | rs6857 | T | C | 0.102 | 0.310 | 0.097 | 0.056 | 0.006 | 1.50E-20 | UTR3 | *NECTIN2* | NM_001042724:c.*618C>T |
| 19 | 44923868 | rs111789331 | A | T | 0.080 | 0.253 | 0.075 | 0.058 | 0.007 | 4.80E-18 | intergenic | *APOC1;APOC1P1* | dist=4524;dist=2935 |
| 19 | 44892652 | rs34404554 | G | C | 0.096 | 0.282 | 0.092 | 0.053 | 0.006 | 2.00E-17 | intronic | *TOMM40* | . |
| 19 | 44892887 | rs11556505 | T | C | 0.097 | 0.282 | 0.092 | 0.053 | 0.006 | 2.10E-17 | exonic | *TOMM40* | . |
| 19 | 44891079 | rs71352238 | C | T | 0.093 | 0.276 | 0.089 | 0.053 | 0.006 | 2.90E-17 | upstream | *TOMM40* | dist=141 |
| 19 | 44892362 | rs2075650 | G | A | 0.097 | 0.282 | 0.092 | 0.052 | 0.006 | 2.90E-17 | intronic | *TOMM40* | . |
| 19 | 44917997 | rs12721046 | A | G | 0.079 | 0.247 | 0.075 | 0.057 | 0.007 | 3.00E-17 | intronic | *APOC1* | . |
| 19 | 44924977 | rs66626994 | A | G | 0.080 | 0.247 | 0.076 | 0.056 | 0.007 | 1.00E-16 | intergenic | *APOC1;APOC1P1* | dist=5633;dist=1826 |
| 19 | 44884202 | rs12972156 | G | C | 0.098 | 0.276 | 0.094 | 0.050 | 0.006 | 4.60E-16 | intronic | *NECTIN2* | . |
| 19 | 44884339 | rs12972970 | A | G | 0.098 | 0.276 | 0.094 | 0.050 | 0.006 | 4.60E-16 | intronic | *NECTIN2* | . |
| 19 | 44883210 | rs142042446 | GTAA | G | 0.098 | 0.276 | 0.094 | 0.050 | 0.006 | 5.10E-16 | intronic | *NECTIN2* | . |
| 19 | 44884873 | rs34342646 | A | G | 0.098 | 0.276 | 0.094 | 0.049 | 0.006 | 6.60E-16 | intronic | *NECTIN2* | . |
| 19 | 44887076 | rs283815 | G | A | 0.170 | 0.391 | 0.164 | 0.037 | 0.005 | 1.10E-14 | intronic | *NECTIN2* | . |
| 19 | 44903416 | rs10119 | A | G | 0.193 | 0.414 | 0.188 | 0.034 | 0.005 | 8.80E-14 | UTR3 | *TOMM40* | NM_001128916:c.*247G>A;NM_006114:c.*247G>A;NM_001128917:c.*247G>A |
| 19 | 44885243 | rs283811 | G | A | 0.172 | 0.385 | 0.166 | 0.036 | 0.005 | 9.10E-14 | intronic | *NECTIN2* | . |
| 19 | 44891712 | rs184017 | G | T | 0.169 | 0.379 | 0.164 | 0.035 | 0.005 | 2.60E-13 | intronic | *TOMM40* | . |
| 19 | 44892962 | rs157582 | T | C | 0.166 | 0.374 | 0.161 | 0.036 | 0.005 | 3.00E-13 | intronic | *TOMM40* | . |
| 19 | 44892457 | rs157581 | C | T | 0.171 | 0.379 | 0.165 | 0.035 | 0.005 | 5.10E-13 | exonic | *TOMM40* | . |
| 19 | 44892587 | rs34095326 | A | G | 0.055 | 0.172 | 0.052 | 0.055 | 0.008 | 4.60E-12 | intronic | *TOMM40* | . |
| 19 | 44913484 | rs438811 | T | C | 0.156 | 0.345 | 0.151 | 0.034 | 0.005 | 6.20E-12 | upstream | *APOC1* | dist=841 |
| 19 | 44912921 | rs483082 | T | G | 0.156 | 0.339 | 0.151 | 0.033 | 0.005 | 2.30E-11 | intergenic | *APOE;APOC1* | dist=3526;dist=1404 |
| 19 | 44915533 | rs5117 | C | T | 0.150 | 0.328 | 0.146 | 0.034 | 0.005 | 3.60E-11 | intronic | *APOC1* | . |
| 19 | 44893408 | rs59007384 | T | G | 0.158 | 0.333 | 0.154 | 0.031 | 0.005 | 4.60E-10 | intronic | *TOMM40* | . |
| 19 | 44910319 | rs75627662 | T | C | 0.135 | 0.276 | 0.131 | 0.030 | 0.005 | 2.30E-08 | downstream | *APOE* | dist=924 |

**Table 3.** Top 15 regions obtained in the SKAT analysis, including both rare and common variants.P-values were adjusted with the Bonferroni method, accounting for the number of regions tested (n = 30,031; p < 1.67E-06). BHP: Bonferroni adjusted P.

| **Chr** | **start position** | **end position** | **SNPs** | **P** | **BHP** | **Symbol** |
|---|---|---|---|---|---|---|
| 19 | 44846175 | 44889223 | 38 | 7.0E-18 | 2.1E-13 | *NECTIN2* |
| 19 | 44905791 | 44909393 | 63 | 8.0E-13 | 2.4E-08 | *APOE* |
| 19 | 44914247 | 44919349 | 63 | 8.0E-13 | 2.4E-08 | *APOC1* |
| 19 | 44890569 | 44903689 | 84 | 1.9E-07 | 5.8E-03 | *TOMM40* |
| 11 | 63624087 | 63671921 | 60 | 5.1E-05 | 1.000 | *ATL3* |
| 17 | 82217934 | 82261129 | 175 | 5.7E-05 | 1.000 | *SLC16A3* |
| 17 | 82236668 | 82236728 | 175 | 5.7E-05 | 1.000 | *MIR6787* |
| 17 | 82239023 | 82273700 | 175 | 5.7E-05 | 1.000 | *CSNK1D* |
| 5 | 104063083 | 104430245 | 98 | 5.9E-05 | 1.000 | *NIHCOLE* |
| 5 | 104780288 | 104780389 | 98 | 5.9E-05 | 1.000 | *RNU6-334P* |
| 5 | 105099473 | 105100098 | 98 | 5.9E-05 | 1.000 | *RAB9BP1* |
| 19 | 1039997 | 1065572 | 146 | 6.3E-05 | 1.000 | *ABCA7* |
| 11 | 63624087 | 63671921 | 121 | 6.4E-05 | 1.000 | *ATL3* |
| 11 | 63632233 | 63632720 | 121 | 6.4E-05 | 1.000 | *IMMP1LP1* |
| 8 | 43140464 | 43202855 | 102 | 6.5E-05 | 1.000 | *HGSNAT* |

**Figure 2:**
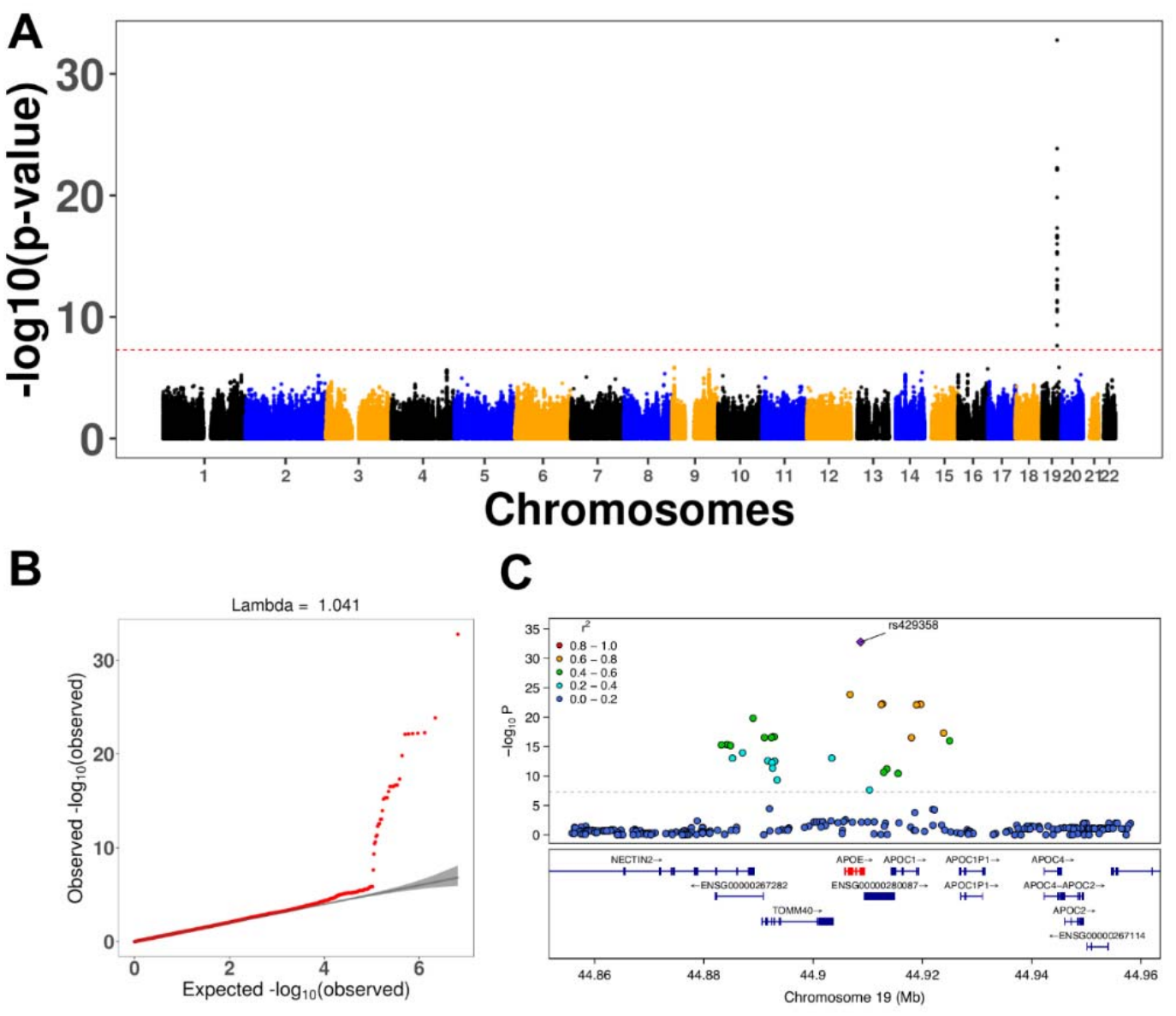
GWAS results. (A) Manhattan plot with significance level (dashed line) at p < 5.0E-08. (B). qq-plot showing no significant inflation. (C) Regional plot showing the details of the APOE region. LD information related to the top SNP rs429358 was estimated from our GWAS dataset using PLINK.

**Figure 3:**
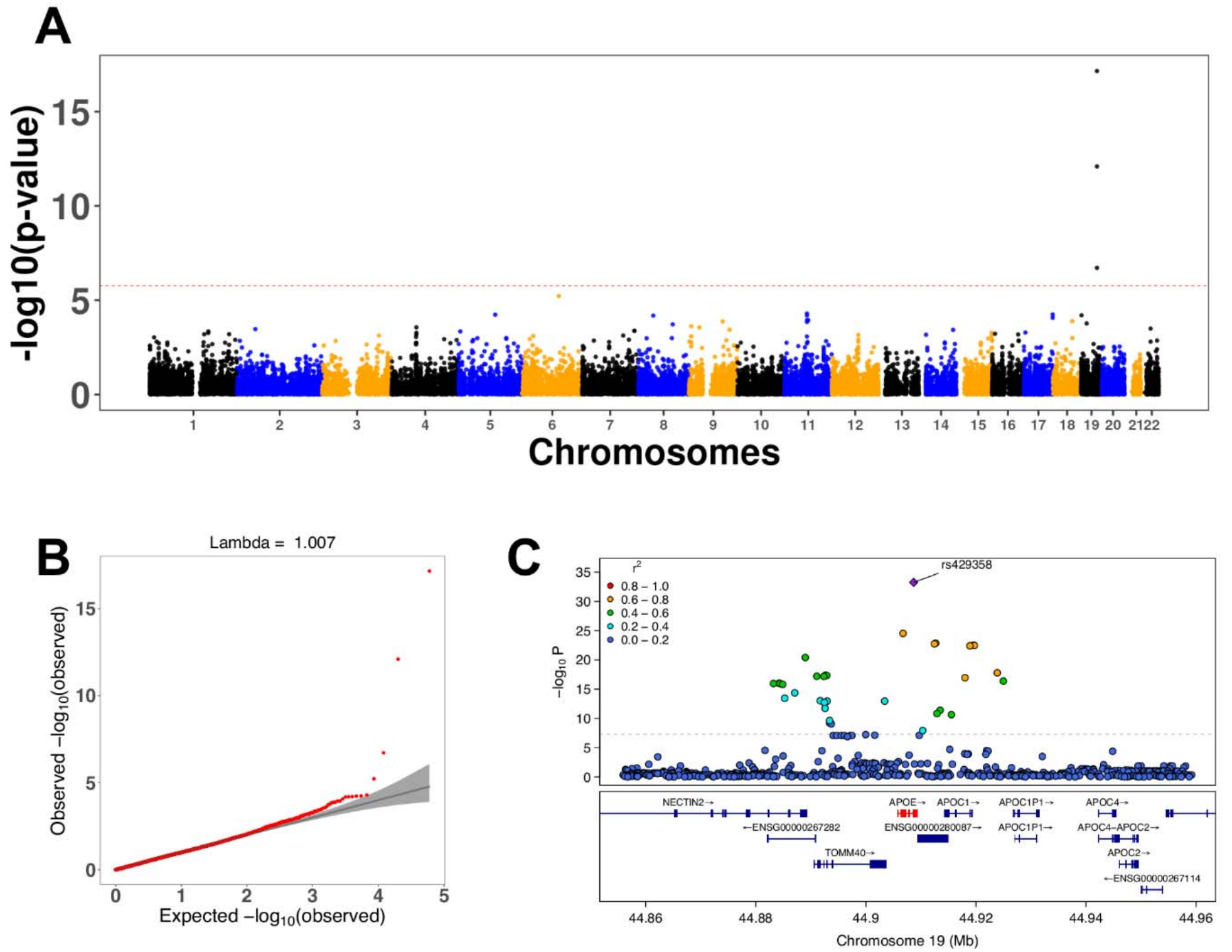
SKAT results including 30,031 regions defining genes based on overlapping transcripts. (A) Manhattan plot with significance level at p < 1.67E-06. (B) qq-plot showing absence of significant inflation. (C) Regional plot showing the BOLT-GWAS p-values for the significant region including variants with MAF.

We calculated a PRS using the GWAS results from Lambert et al.^21^ to assess AD-related risk difference which may explain the anticipate phenotype in EOAD cohort using genes already associated with AD^21^. We selected 3,799 SNPs to calculate PRS for three cohorts: EOAD, TGEN2-AD, and the control cohort TGEN2-ND. The AD-PRS indicates a significantly higher AD risk in the LOAD (TGEN2-AD; *p* = 1.27E-63) and EOAD cohorts (*p* = 1.5E-10) compared to the TGEN2-ND group, with mean AD risk scores of 57.5, 40.1, and 25.1, respectively (***Table 4; Figure 4***). The increased AD risk is statistically significant in the LOAD-affected group, although the EOAD cohort also includes a subset of individuals with notably elevated risk scores (***Figure 4A***).

**Table 4.** Polygenic risk scores across cohorts.

| Cohort | Mean | p-value | t | df | 95% CI |
|---|---|---|---|---|---|
| EOAD | 40.1 |  |  |  |  |
| TGEN2-AD | 57.5 |  |  |  |  |
| TGEN2-ND | 25.1 |  |  |  |  |
| EOAD vs TGEN2-AD |  | 6.68E-05 | 3.52 | 118.2 | 6.56 - 23.42 |
| EOAD vs TGEN2-ND |  | 6.00E-04 | 4.15 | 110.4 | 9.05 - 25.63 |
| TGEN2-AD vs TGEN2-ND |  | 2.20E-16 | 15.11 | 1316.6 | 28.13 - 36.53 |

**Figure 4.**
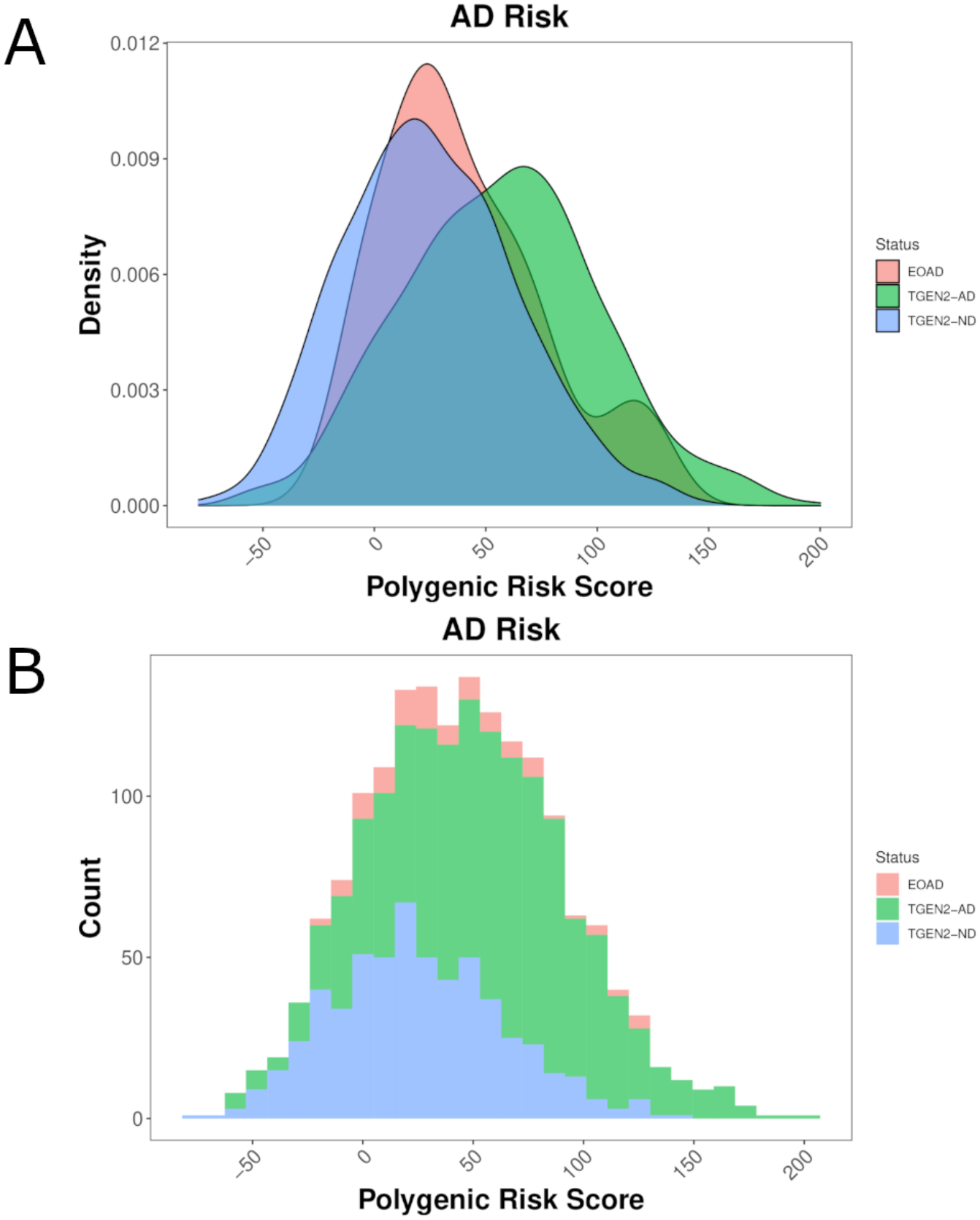
PRS distribution across cohorts. (A) Density histogram of polygenic risk scores stratified by cohort. (B) Frequency histogram of polygenic risk scores showing counts per cohort.

To investigate potential missing genetic risk in EOAD, we compared the distribution of rare (MAF < 0.1%) and singleton variants between EOAD and cognitively normal controls across ten functional consequence categories (***Table S2***). The frequencies of all variant types were highly similar between groups. Missense variants were the most represented class, accounting for over 60% of rare variants and nearly 50% of singleton variants in both EOAD and control samples (***Table S2***). No enrichment was observed for any functional category, including missense as well as putative loss-of-function variants, in either rare or singleton variant sets. These results suggest that sporadic EOAD does not show an increased burden of rare or private functional variants.

## DISCUSSION

Approximately 5% of EOAD cases are explained by recurrent, high-impact familial mutations with clear segregation patterns^2,3^. However, the vast majority of patients lack identifiable variants that can fully account for the early age at onset or inheritance patterns^27^. While rare and private variants are increasingly recognized as potential contributors to disease risk, their interpretation remains challenging. Most are missense variants of uncertain significance, and robust molecular validation is often lacking. Additionally, current studies mainly focus on small variants (<50 bp) captured by exome-based approaches, which cover only a small fraction of the genome and exclude regulatory and structural variants.

To explore the genetic landscape of EOAD, we analyzed WES data from 87 sporadic EOAD patients, 585 late-onset AD (TGEN2-AD) and 1,014 non-demented controls (TGEN2-ND). First, we assessed the distribution of the main genomic risk factor, APOE ε4. The risk allele was significantly more frequent in the EOAD cohort (*p* = 1.54e-10), and in the LOAD cohort (*p* = 1.23e-63) when compared to controls. The APOE ε4 allele demonstrated higher frequency in the LOAD cohort when compared with the EOAD cohort (*p* = 0.0298). Finally, no significant difference was observed in the distribution of APOE ε4 homozygotes between EOAD and LOAD (*p* = 0.8406). While ε4 is a well-established risk factor in LOAD, its role in EOAD remains debated, with some studies suggesting a greater impact in woman^28,29^. Notably, the ε4 allele is neither necessary nor sufficient to cause AD. In LOAD, it influences age at onset in a dose-dependent manner^2^. In autosomal dominant EOAD (*APP, PSEN1*, and *PSEN2*), age at onset varies widely despite identical APOE4 genotype, consistent with the semidominant inheritance of a moderately penetrant gene^2^. Our results here suggest that the e4 allele is a significant genetic risk factor for EOAD even though this influence may prove to be less so than for LOAD.

To investigate the contribution of common variants, we perform a GWAS analysis on 87 EOAD and 3,487 not-demented controls (ADSP-CTL). This analysis confirmed the central role of the *APOE* locus, with a significant GWAS signal involving 31 SNPs on chromosome 19 (p < 5.0e-08), including *APOE, APOC1, NECTIN2*, and *TOMM40*. The strongest association was observed for rs429358 within *APOE* (*p* = 9.8e-28) (***Figure 2; Table 2***), which, along with rs7412, determines the ε2/ε3/ε4 isoforms. The C allele at rs429358 encode the ε4 isoform, which confers the highest genetic risk factor for AD, particularly for LOAD^30^. To our knowledge, this represents the first GWAS on sporadic EOAD.

SKAT analysis revealed significant enrichment in four genes within the APOE locus in EOAD compared to controls. *NECTIN2* and *APOE* showed the strongest association after Bonferroni correction (*p* = 2.1e-13 and 2.4e-8, respectively), followed by *APOEC1* and *TOMM40* (***Figure 3; Table 3***). Although the APOE ε4 allele has been extensively studied as a modifier in both EOAD and LOAD, its influence on clinical phenotype in EOAD remains less well defined. Some reports highlights a stronger effect in autosomal dominant and LOAD forms, while others suggest differences related to sporadic versus familial origin, or even sex-specific effects^28,31–35^. The concordance of both common and rare variant signals at the *APOE* locus may suggest a strong locus-specific effect, though contributions from other genomic regions may remain undetected due to the limitations of exome sequencing and SNP-based approaches.

We calculated PRS across three different cohorts: EOAD, neuropathological confirmed LOAD (TGEN2-AD) and non-demented controls (TGEN2-ND). PRS were significantly lower in EOAD than in LOAD (*p* = 6.68e-05), but higher than in controls (*p* = 6e-04), suggesting an intermediate polygenic burden and/or the presence of distinct risk factors not captured by this approach. These results are consistent with previous studies showing similar risk factors, and neuropathological features between mendelian EOAD and LOAD^15,36,37^. Notably, PRS distribution within the EOAD cohort revealed a subgroup with higher scores, supporting substantial genetic heterogeneity, both across AD subtypes and within sporadic EOAD itself (***Figure 4***). However, the discrepancy in genetic risk between EOAD and LOAD may reflect inherent limitations of current PRS models, highlighting the need for refined approaches capable of capture a broader spectrum of risk factors, including complex genomic variants^7^. Indeed, these PRS models are based on LOAD and on European ancestry, limiting their ability to capture genetic risk factors specific for EOAD or for other ancestries. Previous studies have also show that PRS perform better when adjasted for gender, age and APOE4 genotype^38^.

In conclusion, we further investigated the contribution of rare variants in a curated list of 83 genes previously associated with EOAD and LOAD. Interestingly, we did not observe a significant enrichment of rare or singleton variants within these genes, suggesting that such variants may not contribute substantially to disease risk in our cohort (***Table S2***). This finding contrasts with some studies reporting an enrichment of rare variants in genes like *TREM2, SORL1* and *ABCA7* in EOAD cohorts^39–41^. This discrepancy underscores the genomic complexity of EOAD and may reflect differences in patient selection criteria, as well as analytical approaches and variant filtering strategies aimed at identifying potentially pathogenic rare variants^39–41^. Despite increasing efforts, the literature and our work still present key limitations, including the incomplete or missing analysis of noncoding regions, limited investigation of complex variants such as CNVs and structural rearrangements, and a lack of integrative approaches that combine these data with SNP-based models. While genetic factors account for approximately 30% of the risk of developing Alzheimer’s disease, epigenetic regulation may exert a broader influence by modulating multiple genes and pathways. Moreover, neurodegeneration should be viewed within a systemic framework, as factors such as gene–environment interactions, lifestyle, and the gut microbiome can affect brain aging, even indirectly. Additionally, the predominance of studies conducted in cohorts of European ancestry further restricts the generalizability of findings. A deeper understanding of the genetic architecture and etiological complexity of EOAD is essential to refine individual risk prediction, uncover disease mechanisms, and ultimately improve clinical management, enhancing diagnostic and prognostic precision and enabling the development of novel therapeutic strategies, which remains a critical priority across all forms of AD.

## Data Availability

All data generated or analyzed during this study are included in the manuscript and its supplementary materials.

## Statements & Declarations Ethics approval

The study was conducted in accordance with relevant institutional ethical guidelines and approvals.

Baylor Scott & White samples were obtained under Baylor IRB approval #017-176, and Mayo Clinic Arizona samples under WCG Protocol #20120612. Informed consent was obtained from all participants or their legally authorized representatives, as applicable.

## Acknowledgments

The ADSP-CTL cohort data was obtained from the NIH-designated database of Genotypes and Phenotypes (dbGaP) under accession number phs000572.v7.p4. We acknowledge the contributions of the Principal Investigators Richard A. Gibbs, PhD (Human Genome Sequencing Center, Baylor College of Medicine, Houston, TX, USA), Eric S. Lander, PhD (Broad Institute, Boston, MA, USA), Richard K. Wilson, PhD (The Genome Institute, Washington University, St. Louis, MO, USA), Gerard D. Schellenberg, PhD (Alzheimer’s Disease Genetics Consortium, University of Pennsylvania, Philadelphia, PA, USA), and Sudha Seshadri, MD (Cohorts for Heart and Aging Research in Genomic Epidemiology Consortium, Boston University, Boston, MA, USA). The generation of data for phs000572.v7.p4 was supported by the following funding sources: U24 AG041689, U01 AG032984, R01 AG033193, U24 AG021886, U54HG003079, U54HG003273, and U54HG003076, funded by the National Institutes of Health (NIH), including the National Institute on Aging (NIA) and the National Human Genome Research Institute (NHGRI). We gratefully acknowledge the investigators who submitted the data and the funding organizations that made this study possible.

## Author contributions

Clinical assessments were performed by CRP, RC, BW. Neuropathological assessments were performed by TB and GS. Sample preparation, data collection and analysis were performed by BA, NM, FE and PIS. The first draft of the manuscript was written by FE and all authors participated in the writing or revision of this manuscript: FE, LN, MN, AB, ISP, MJH, and CRP. All authors read and approved the final manuscript.

## Figure Legends

**Table 1**. Demographic and clinical characteristic of AD patients and control samples enrolled in the study.

**Figure 1**. APOE4 allele distribution across the cohorts.

**Figure 2**. GWAS results. (A) Manhattan plot with significance level (dashed line) at p < 5.0E-08. (B). qq-plot showing no significant inflation. (C) Regional plot showing the details of the APOE region. LD information related to the top SNP rs429358 was estimated from our GWAS dataset using PLINK.

**Table 2**. Significant SNPs (p < 5.0E-08) obtained in the GWAS comparison EAOD (n = 87) and ADSP-CTL (n = 3487). A1: effect allele; A2; other allele.

**Figure 3**. SKAT results including 30,031 regions defining genes based on overlapping transcripts. (A) Manhattan plot with significance level at p < 1.67E-06. (B) qq-plot showing absence of significant inflation. (C) Regional plot showing the BOLT-GWAS p-values for the significant region including variants with MAF >= 0.1%. LD information related to the top SNP rs429358 was estimated from our GWAS dataset using PLINK.

**Table 3**. Top 15 regions obtained in the SKAT analysis, including both rare and common variants. P-values were adjusted with the Bonferroni method, accounting for the number of regions tested (n = 30,031; p < 1.67E-06). BHP: Bonferroni adjusted P.

**Figure 4**. PRS distribution across cohorts. (A) Density histogram of polygenic risk scores stratified by cohort. (B) Frequency histogram of polygenic risk scores showing counts per cohort.

**Table 4**. Polygenic risk scores (PRS) across cohorts, with statistical comparisons of PRS between cohorts.

**Table S1**. Curated gene list associated with early-onset and late-onset Alzheimer’s Disease.

**Table S2**. Variant type frequencies for rare and singleton variants in EOAD and controls.

## Bibliography

1. Van Der Flier WM, De Vugt ME, Smets EMA, Blom M, Teunissen CE. Towards a future where Alzheimer’s disease pathology is stopped before the onset of dementia. Nat Aging. 2023 May 18;3(5):494–505. doi:10.1038/s43587-023-00404-2

2. Reitz C, Rogaeva E, Beecham GW. Late-onset vs nonmendelian early-onset Alzheimer disease: A distinction without a difference? Neurol Genet. 2020 Oct;6(5):e512. doi:10.1212/NXG.0000000000000512 PubMed PMID: 33225065; PubMed Central PMCID: PMC7673282.

3. Hoogmartens J, Cacace R, Van Broeckhoven C. Insight into the genetic etiology of Alzheimer’s disease: A comprehensive review of the role of rare variants. Alz & Dem Diag Ass & Dis Mo. 2021 Jan;13(1):e12155. doi:10.1002/dad2.12155

4. Sirkis DW, Bonham LW, Johnson TP, La Joie R, Yokoyama JS. Dissecting the clinical heterogeneity of early-onset Alzheimer’s disease. Mol Psychiatry. 2022 Jun;27(6):2674–88. doi:10.1038/s41380-022-01531-9

5. Stoychev KR, Stoimenova-Popova M, Chumpalova P, Ilieva L, Swamad M, Kamburova-Martinova Z. A Clinical Case of Patient Carrying Rare Pathological PSEN1 Gene Mutation (L424V) Demonstrates the Phenotypic Heterogenity of Early Onset Familial AD. Front Psychiatry. 2019 Dec 11;10:857. doi:10.3389/fpsyt.2019.00857

6. ARUK Consortium, GERAD/PERADES, CHARGE, ADGC, EADI, Sims R, Van Der Lee SJ, Naj AC, Bellenguez C, et al. Rare coding variants in PLCG2, ABI3, and TREM2 implicate microglial-mediated innate immunity in Alzheimer’s disease. Nat Genet. 2017 Sep;49(9):1373–84. doi:10.1038/ng.3916

7. Han L, Zhao X, Benton ML, Perumal T, Collins RL, Hoffman GE, et al. Functional annotation of rare structural variation in the human brain. Nat Commun. 2020 Jun 12;11(1):2990. doi:10.1038/s41467-020-16736-1 PubMed PMID: 32533064; PubMed Central PMCID: PMC7293301.

8. Lek M, Karczewski KJ, Minikel EV, Samocha KE, Banks E, Fennell T, et al. Analysis of protein-coding genetic variation in 60,706 humans. Nature. 2016;536(7616):285–91. doi:10.1038/nature19057 PubMed PMID: 27535533 PMCID: PMC5018207.

9. Blauwendraat C, Nalls MA, Singleton AB. The genetic architecture of Parkinson’s disease. The Lancet Neurology. 2020 Feb;19(2):170–8. doi:10.1016/S1474-4422(19)30287-X

10. Li T, Ferraro N, Strober BJ, Aguet F, Kasela S, Arvanitis M, et al. The functional impact of rare variation across the regulatory cascade. Cell Genomics. 2023 Oct;3(10):100401. doi:10.1016/j.xgen.2023.100401

11. Cirulli ET, White S, Read RW, Elhanan G, Metcalf WJ, Tanudjaja F, et al. Genome-wide rare variant analysis for thousands of phenotypes in over 70,000 exomes from two cohorts. Nat Commun. 2020 Jan 28;11(1):542. doi:10.1038/s41467-020-14288-y

12. Vialle RA, de Paiva Lopes K, Bennett DA, Crary JF, Raj T. Integrating whole-genome sequencing with multi-omic data reveals the impact of structural variants on gene regulation in the human brain. Nat Neurosci. 2022 Apr;25(4):504–14. doi:10.1038/s41593-022-01031-7 PubMed PMID: 35288716; PubMed Central PMCID: PMC9245608.

13. Miller MB, Reed HC, Walsh CA. Brain Somatic Mutation in Aging and Alzheimer’s Disease. Annu Rev Genom Hum Genet. 2021 Aug 31;22(1):239–56. doi:10.1146/annurev-genom-121520-081242

14. Esai Selvan M, Onel K, Gnjatic S, Klein RJ, Gümüş ZH. Germline rare deleterious variant load alters cancer risk, age of onset and tumor characteristics. npj Precis Onc. 2023 Jan 27;7(1):13. doi:10.1038/s41698-023-00354-3

15. Shen Y, Timsina J, Heo G, Beric A, Ali M, Wang C, et al. CSF proteomics identifies early changes in autosomal dominant Alzheimer’s disease. Cell. 2024 Sep;S0092867424009784. doi:10.1016/j.cell.2024.08.049

16. Jin SC, Benitez BA, Karch CM, Cooper B, Skorupa T, Carrell D, et al. Coding variants in TREM2 increase risk for Alzheimer’s disease. Hum Mol Genet. 2014 Nov 1;23(21):5838–46. doi:10.1093/hmg/ddu277 PubMed PMID: 24899047; PubMed Central PMCID: PMC4189899.

17. Chang CC, Chow CC, Tellier LC, Vattikuti S, Purcell SM, Lee JJ. Second-generation PLINK: rising to the challenge of larger and richer datasets. Gigascience. 2015 Dec 1;4(1):s13742-015-0047–8. doi:10.1186/s13742-015-0047-8

18. Loh PR, Tucker G, Bulik-Sullivan BK, Vilhjálmsson BJ, Finucane HK, Salem RM, et al. Efficient Bayesian mixed-model analysis increases association power in large cohorts. Nat Genet. 2015 Mar;47(3):284–90. doi:10.1038/ng.3190 PubMed PMID: 25642633; PubMed Central PMCID: PMC4342297.

19. Gogarten SM, Sofer T, Chen H, Yu C, Brody JA, Thornton TA, et al. Genetic association testing using the GENESIS R/Bioconductor package. Bioinformatics. 2019 Dec 15;35(24):5346–8. doi:10.1093/bioinformatics/btz567 PubMed PMID: 31329242; PubMed Central PMCID: PMC7904076.

20. Marees AT, de Kluiver H, Stringer S, Vorspan F, Curis E, Marie-Claire C, et al. A tutorial on conducting genome-wide association studies: Quality control and statistical analysis. Int J Methods Psychiatr Res. 2018 Jun;27(2):e1608. doi:10.1002/mpr.1608 PubMed PMID: 29484742; PubMed Central PMCID: PMC6001694.

21. Lambert JC, Ibrahim-Verbaas CA, Harold D, Naj AC, Sims R, Bellenguez C, et al. Meta-analysis of 74,046 individuals identifies 11 new susceptibility loci for Alzheimer’s disease. Nat Genet. 2013 Dec;45(12):1452–8. doi:10.1038/ng.2802 PubMed PMID: 24162737; PubMed Central PMCID: PMC3896259.

22. Corneveaux JJ, Myers AJ, Allen AN, Pruzin JJ, Ramirez M, Engel A, et al. Association of CR1, CLU and PICALM with Alzheimer’s disease in a cohort of clinically characterized and neuropathologically verified individuals. Hum Mol Genet. 2010 Aug 15;19(16):3295–301. doi:10.1093/hmg/ddq221 PubMed PMID: 20534741; PubMed Central PMCID: PMC2908469.

23. Cho H, Kim YE, Chae W, Kim KW, Kim JW, Kim HJ, et al. Distribution and clinical impact of apolipoprotein E4 in subjective memory impairment and early mild cognitive impairment. Sci Rep. 2020 Aug 7;10(1):13365. doi:10.1038/s41598-020-69603-w

24. Troutwine BR, Hamid L, Lysaker CR, Strope TA, Wilkins HM. Apolipoprotein E and Alzheimer’s disease. Acta Pharmaceutica Sinica B. 2022 Feb;12(2):496–510. doi:10.1016/j.apsb.2021.10.002

25. Block AD, Castro MR, Hansen A, Strong C, Hanes E, Schomaker ML, et al. APOE ε4 Allele Frequency Among Cognitively Healthy Members of an Ojibwe Tribal Nation. Neurol Genet. 2024 Apr;10(2):e200130. doi:10.1212/NXG.0000000000200130

26. Katsumata Y, Wu X, Aung KZ, Fardo DW, Woodworth DC, Sajjadi SA, et al. Pure LATE-NC: Frequency, clinical impact, and the importance of considering APOE genotype when assessing this and other subtypes of non-Alzheimer’s pathologies. Acta Neuropathol. 2024 Nov 15;148(1):66. doi:10.1007/s00401-024-02821-y

27. Uddin MdS, Hasana S, Hossain MdF, Islam MdS, Behl T, Perveen A, et al. Molecular Genetics of Early- and Late-Onset Alzheimer’s Disease. CGT. 2021 Feb 9;21(1):43–52. doi:10.2174/1566523220666201123112822

28. Nemes S, Logan PE, Manchella MK, Mundada NS, La Joie R, Polsinelli AJ, et al. Sex and APOE ε4 carrier effects on atrophy, amyloid PET, and tau PET burden in early-onset Alzheimer’s disease. Alzheimer’s & Dementia. 2023 Nov;19(S9). doi:10.1002/alz.13403

29. Polsinelli AJ, Logan PE, Lane KA, Manchella MK, Nemes S, Sanjay AB, et al. APOE ε4 carrier status and sex differentiate rates of cognitive decline in early- and late-onset Alzheimer’s disease. Alzheimer’s & Dementia. 2023 May;19(5):1983–93. doi:10.1002/alz.12831

30. Blanchard JW, Akay LA, Davila-Velderrain J, von Maydell D, Mathys H, Davidson SM, et al. APOE4 impairs myelination via cholesterol dysregulation in oligodendrocytes. Nature. 2022 Nov;611(7937):769–79. doi:10.1038/s41586-022-05439-w PubMed PMID: 36385529; PubMed Central PMCID: PMC9870060.

31. Van Der Flier WM, Pijnenburg YA, Fox NC, Scheltens P. Early-onset versus late-onset Alzheimer’s disease: the case of the missing APOE [4 allele. The Lancet Neurology. 2011 Mar;10(3):280–8. doi:10.1016/S1474-4422(10)70306-9

32. Van Der Vlies AE, Koedam ELGE, Pijnenburg YAL, Twisk JWR, Scheltens P, Van Der Flier WM. Most rapid cognitive decline in APOE ε4 negative Alzheimer’s disease with early onset. Psychol Med. 2009 Nov;39(11):1907–11. doi:10.1017/S0033291709005492

33. Houlden H, Crook R, Backhovens H, Prihar G, Baker M, Hutton M, et al. ApoE genotype is a risk factor in nonpresenilin early-onset alzheimer’s disease families. Am J Med Genet. 1998 Feb 7;81(1):117–21. doi:10.1002/(SICI)1096-8628(19980207)81:1<117::AID-AJMG19>3.0.CO;2-M

34. Balasa M, Gelpi E, Antonell A, Rey MJ, Sánchez-Valle R, Molinuevo JL, et al. Clinical features and APOE genotype of pathologically proven early-onset Alzheimer disease. Neurology. 2011 May 17;76(20):1720–5. doi:10.1212/WNL.0b013e31821a44dd

35. Kaur G, Poljak A, Braidy N, Crawford JD, Lo J, Sachdev PS. Fluid Biomarkers and APOE Status of Early Onset Alzheimer’s Disease Variants: A Systematic Review and Meta-Analysis. Journal of Alzheimer’s Disease. 2020 Jun 2;75(3):827–43. doi:10.3233/JAD-200052

36. Lippa CF, Saunders AM, Smith TW, Swearer JM, Drachman DA, Ghetti B, et al. Familial and sporadic Alzheimer’s disease: Neuropathology cannot exclude a final common pathway. Neurology. 1996 Feb;46(2):406–12. doi:10.1212/WNL.46.2.406

37. Cruchaga C, Del-Aguila JL, Saef B, Black K, Fernandez MV, Budde J, et al. Polygenic risk score of sporadic late-onset Alzheimer’s disease reveals a shared architecture with the familial and early-onset forms. Alzheimer’s & Dementia. 2018 Feb;14(2):205–14. doi:10.1016/j.jalz.2017.08.013

38. Escott-Price V, Shoai M, Pither R, Williams J, Hardy J. Polygenic score prediction captures nearly all common genetic risk for Alzheimer’s disease. Neurobiology of Aging. 2017 Jan;49:214.e7-214.e11. doi:10.1016/j.neurobiolaging.2016.07.018

39. Holstege H, Hulsman M, Charbonnier C, Grenier-Boley B, Quenez O, Grozeva D, et al. Exome sequencing identifies rare damaging variants in ATP8B4 and ABCA1 as risk factors for Alzheimer’s disease. Nat Genet. 2022 Dec;54(12):1786–94. doi:10.1038/s41588-022-01208-7

40. Bellenguez C, Charbonnier C, Grenier-Boley B, Quenez O, Le Guennec K, Nicolas G, et al. Contribution to Alzheimer’s disease risk of rare variants in TREM2, SORL1, and ABCA7 in 1779 cases and 1273 controls. Neurobiology of Aging. 2017 Nov;59:220.e1-220.e9. doi:10.1016/j.neurobiolaging.2017.07.001

41. Lacour M, Quenez O, Rovelet-Lecrux A, Salomon B, Rousseau S, Richard AC, et al. Causative Mutations and Genetic Risk Factors in Sporadic Early Onset Alzheimer’s Disease Before 51 Years. Journal of Alzheimer’s Disease. 2019 Sep 3;71(1):227–43. doi:10.3233/JAD-190193

